# Gene-gene interactions protect against Familial Hypercholesterolemia: effect of lost- of-function *PCSK9* variants

**DOI:** 10.64898/2026.03.26.26348145

**Authors:** Sonia Rodríguez-Nóvoa, Pedro Martínez Hernández, Irene Hidalgo Mayoral, Amanda Herranz Cecilia, Neila Rodríguez Roca, Ana Carazo Álvarez, Natividad Gallego Onís, Marta Duque Alcorta, Carmen Rodríguez Jiménez

**Author notes:** **Corresponding authors:** Sonia Rodríguez-Nóvoa, Genetics of Metabolic Diseases Laboratory, Genetics Department, Hospital Universitario La Paz, Paseo de la Castellana 261, 28046, Madrid, Spain, Carmen Rodríguez Jiménez, Genetics of Metabolic Diseases Laboratory, Genetics Department, Hospital Universitario La Paz, Paseo de la Castellana 261, 28046, Madrid, Spain.

## Abstract

**Background:** Familial hypercholesterolemia (FH) is most frequently caused by pathogenic variants in *LDLR*, but phenotypic variability suggests the influence of genetic modifiers.

**Methods:** We investigated a large multigenerational family with FH, combining clinical data, lipid profiles, and genetic analysis with functional studies. *LDLR* and *PCSK9* variants were characterized according to ACMG/ClinGen guidelines. Functional assays in CHO-ldlA7 cells assessed *LDLR* activity, while plasma PCSK9 levels were quantified by ELISA.

**Results:** *LDLR* c.2479G>A variant was associated with FH in the family. The presence of loss of function *c.137G>T* and c.2023del variants at *PCSK9* appears to mitigate the effect of the *LDLR* variant.

**Conclusions:** This study provides evidence that *PCSK9* variants can counteract the deletereous effect of a *LDLR* variant associated with FH. These findings highlight the importance of gene–gene interactions in the clinical variability of FH and their potential implications for precision medicine.

## Introduction

Familial hypercholesterolemia (FH) is an autosomal disorder of lipid metabolism characterized by elevated levels of total cholesterol (TC) and low-density lipoprotein cholesterol (LDL-C), which predispose affected individuals to premature atherosclerotic cardiovascular disease (ASCVD). If untreated, FH is associated with an increased risk of myocardial infarction and cardiovascular mortality, frequently manifesting decades earlier than in the general population. Despite being a monogenic condition, it remains underdiagnosed and undertreated worldwide, representing a significant public health challenge(1) (2). The estimated prevalence of heterozygous FH is approximately 1 in 200–250 individuals, making it one of the most common inherited metabolic disorders, whereas homozygous FH is considerably rarer, affecting about 1 in 160,000–300,000 individuals(3).

Clinical diagnosis of FH is based on standardized criteria such as the Dutch Lipid Clinic Network (DLCN) score, which incorporates lipid values, clinical signs (xanthomas, corneal arcus), family history of hypercholesterolemia or premature ASCVD, and the presence of a pathogenic genetic variant(4). Nevertheless, the FH phenotype can be variable and may be subtle or even absent in childhood or early adulthood. Early diagnosis is essential, as timely initiation of statins and other lipid-lowering therapies can restore near-normal life expectancy for most. Consequently, cascade genetic screening of relatives is recommended once an index case is identified(5). The incorporation of genetic testing into FH diagnostic workflows significantly increases the detection rate, facilitates the identification of affected relatives, and enables more precise cardiovascular risk stratification.

Approximately 90% of genetically confirmed FH cases are caused by pathogenic loss-of-function (LOF) variants in the low-density lipoprotein receptor gene (*LDLR*, MIM 606945)(6). Additional monogenic causes include deleterious variants in the apolipoprotein B gene (*APOB*, MIM 107730), that impair the ability of LDL-C particles to bind LDL receptor (LDLR)(7), and gain-of-function (GOF) variants in proprotein convertase subtilisin/kexin type 9 (*PCSK9*, MIM 607786), which accelerate LDLR degradation(8). Rare biallelic LoF variants in LDL receptor adaptor protein 1 (*LDLRAP1*, MIM 605747) cause the autosomal recessive form of FH, frequently characterised by extremely increased LDL-C levels and early onset ASCVD(9).

Although FH is considered a monogenic disease, its clinical expression is highly heterogeneous. Individuals carrying the same pathogenic variant, even within a single family, may exhibit markedly differences in LDL-C levels, age at onset of cardiovascular events, or response to lipid lowering therapy. This variability suggests the influence of genetic, epigenetic, and environmental modifiers. The identification of genetic modifiers is particularly relevant, as they may explain “resilience” or unexpected mild phenotypes in certain carriers of pathogenic variants and may also provide insights into novel therapeutic approaches.

The *PCSK9* gene encodes proprotein convertase subtilisin/kexin type 9 (PCSK9), a secreted serine protease that plays a central role in cholesterol homeostasis. PCSK9 regulates the number of LDLR on the hepatocyte surface by binding to LDLR and targeting it for lysosomal degradation. Consequently, increased PCSK9 activity leads to higher plasma LDL-C levels, whereas reduced activity promotes LDL clearance. Since its identification as an FH gene in 2003(8), PCSK9 has attracted considerable attention both for its role in monogenic disease and as a therapeutic target. Monoclonal antibody inhibitors of PCSK9 are now widely used in clinical practice, and small interfering RNA therapies are emerging as long-acting alternatives.

Remarkably, PCSK9 also represents one of the best-characterized examples of naturally occurring protective genetic variants. Carriers of LOF variants in PCSK9 exhibit lifelong lower LDL-C levels and significantly lower risk of coronary artery disease without apparent adverse effects(10). These observations firmly establish that PCSK9 activity directly influences cardiovascular risk. Furthermore, the coexistence of variants in PCSK9 and other FH-related genes raises the potential for gene–gene interactions that may modulate disease severity. However, reports of such interactions remain limited, and functional validation is seldom available.

Herein, we report a family with FH carrying a controversial *LDLR* variant, and describe functional studies alongside a critical appraisal of the available evidence to discuss its potential pathogenicity. Additionally, we report the effect of a rare LOF PCSK9 genotype and its effect on the phenotypic expression of the disease in this family.

## Patients and Methods

### Patients and ethical approval

The index case was a male in his fifties with a clinical diagnosis of FH according to the Dutch Lipid Clinic Network (DLCN) criteria, who was referred for genetic testing. Following the index case results, cascade genetic testing was offered to all available relatives. All participating family members provided written informed consent for genetic analysis in accordance with the Declaration of Helsinki, and the study was approved by the institutional ethics committee. Clinical data was collected, including lipid profiles, cardiovascular history, and physical examination (xanthomas, corneal arcus).

### Genetic analysis

Peripheral blood samples were collected in EDTA tubes, and genomic DNA was extracted using the QIAamp DNA Blood Mini Kit (Qiagen, Hilden, Germany) following the manufacturer’s instructions. DNA concentration and purity were assessed by NanoDrop™ spectrophotometry and Qubit fluorometry (Thermo Fisher Scientific).

Genetic testing was performed by targeted next-generation sequencing (NGS) using a custom panel of 198 genes related to lipid metabolism. Library preparation and exome enrichment were carried out according to the manufacturer’s protocol (Roche Nimblegen, Madison, WI, USA), and sequencing was performed on a MiSeq system (Illumina, San Diego, CA, USA). For this study, analyses focused on the four major FH genes: *LDLR, APOB, PCSK9*, and *LDLRAP1*.

Bioinformatic analysis: variants were interpreted using in silico pathogenicity predictors (CADD, PolyPhen-2, MutAssessor, FATHMM, VEST), conservation scores (GERP2, PhastCons, PhyloP), and splicing predictors (MaxEntScan, NNSplice, GeneSplicer, Human Splicing Finder). Variants with minor allele frequency (MAF) >1% in population databases (gnomAD, 1000 Genomes) were excluded. BAM files were analyzed using Alamut Visual (SOPHiA Genetics, Lausanne, Switzerland). Variant classification followed ACMG/AMP and ClinGen guidelines(11) (12).

Large rearrangements in *LDLR* were screened by multiplex ligation-dependent probe amplification (MLPA; MRC-Holland, Amsterdam, The Netherlands). Variants of interest and segregation analysis were confirmed by Sanger sequencing using BigDye Terminator v3.1 Cycle Sequencing Kit and analyzed on an ABI 3500 Genetic Analyzer (Applied Biosystems).

### Functional studies of the LDLR variant

#### Cell culture and transfection

LDLR-deficient CHO-ldlA7 cells (kindly provided by Dr. Monty Krieger, Massachusetts Institute of Technology, Cambridge, MA, USA)(13) were maintained in Ham’s F-12 medium (Gibco, Life Technologies, Carlsbad, CA, USA) supplemented with 5% heat-inactivated fetal bovine serum (FBS), 100 U/mL penicillin, 100 µg/mL streptomycin, 2 mmol/L L-glutamine, and 50 mg/mL Normocin (InvivoGen, Toulouse, France). Cells were cultured at 37 °C in a humidified atmosphere with 5% CO_2_ and passaged every 2-3 days.

Human LDLR cDNA constructs were generated using a GFP-tagged plasmid vector (C-GFPSpark®, NM_000527; Sino Biological, Beijing, China). Site-directed mutagenesis was performed (QuikChange Lightning kit; Agilent Technologies, Santa Clara, CA, USA) to introduce the variant of interest. Plasmid integrity and the presence of the intended mutation were confirmed by Sanger sequencing.

Transient transfections were performed in 24-well plates (1.5 × 10^5^ cells/well) using Lipofectamine 3000 (Invitrogen™, Thermo Fisher Scientific, Waltham, MA, USA) according to the manufacturer’s instructions. Cells were incubated for 48 h before downstream analyses.

#### LDLR activity by flow cytometry

LDL was isolated from a healthy donor and labeled with DiI (1,1′-dioctadecyl-3,3,3,3′-tetramethylindocarbocyanine perchlorate; Molecular Probes, Life Technologies). Transfected CHO-ldlA7 cells were incubated with 20 µg/mL DiI-LDL for 4 h at either 4 °C to measures LDL binding, or 37 °C to assess LDL uptake/internalisation. After incubation, cells were washed with PBS-2% BSA and resuspended in PBS with 0.1% DAPI to exclude dead cells. Flow cytometry was performed using a BD FACSCanto™ II cytometer (BD Biosciences, San Jose, CA, USA), acquiring at least 10^4^ events per sample. Experiments were performed in triplicate, and data were anlyzed using FlowJo v10 software.

#### 3D modelling of PCSK9 protein structure

A 3D model of PCSK9 (PDB ID: 2PMW) was obtained from the RCSB (14) protein data bank and PCSK9 mutant structure was generated by SWISS-MODEL (workspace homology modelling using AF-Q8NBP7-F1 PCSK9 model from AlphaFold as a template (15). 3D structures were visualized by UCSF-Chimera (16).

### Quantification of serum PCSK9

Secreted PCSK9 protein levels in culture supernatants were quantified using a commercial enzyme-linked immunosorbent assay (Human PCSK9 ELISA Kit, Cell Biolabs Inc., San Diego, CA, USA) according to the manufacturer’s specifications. Samples were measured in duplicate. Absorbance was read at 450 nm with background correction at 570 nm. PCSK9 concentration were calculated against a standard curve generated with recombinant human PCSK9.

## 3. Results

### 3.1 Clinical characteristics of the proband

The index case was a male patient clinically diagnosed with FH in early adulthood and subsequently followed regularly in a endocrinology outpatient clinic. His maximum recorded total cholesterol and LDL cholesterol levels were 335 mg/dL and 265 mg/dL, respectively. In his forties’, while receiving atorvastatin 10 mg daily and with total cholesterol and LDL cholesterol levels of 113 mg/dL and 50 mg/dL, respectively, he experienced several episodes of retrosternal chest pain radiating to the left shoulder during physical exercise. Coronary angiography revealed a single-vessel obstructive lesion involving the circumflex coronary artery. He was therefore diagnosed with chronic ischemic heart disease manifesting as stable effort angina. Treatment included coronary stent implantation and intensification of lipid-lowering therapy to atorvastatin 40 mg daily. Genetic testing was performed in the patient, and the results are presented below.

### 3.2 Proband’s molecular analysis

Molecular analysis revealed the presence of the following heterozygous variants: *LDLR* (NM_000527.5) c.2479G>A p.(Val827Ile), *PCSK9 (NM_174936.4) c.137G>T* p.(Arg46Leu) and *APOB (NM_000384.3) c.2630C>T* p.(Pro877Leu).

The ***LDLR* c.2479G>A**, consists of a guanine to adenine nucleotide substitution that predicts the replacement of aminoacid valine to isoleucine at position 827 of the protein, located in the C-terminal cytosolic domain of the LDL receptor. It is present at a minor frequency (MAF) of 1,83% in Ashkenazi Jewis exomes-genomes at the Genome Aggregation Database (gnomAD-v4.1.0), four homozygotes reported, and despite its frequency it has been previously reported in the context of FH as a likely pathogenic variant in ClinVar, supporting a molecular diagnosis of FH in our patient. However, subsequent submissions have led to variable reclassification of this variant, ranging from likely pathogenic to variant of uncertain significance (VUS). According to the most recent ACMG/ClinGen FH-specific LDLR curation guidelines: variant segregates with FH phenotype (PP1) and in silico predictors suggest a deleterious impact (PP3).

The ***PCSK9* c.137G>T** consists of a guanine to timine nucleotide substitution that predicts the replacement of amino acid arginine to leucine at position 46 of the protein, located in the prodomain of PCSK9. It is present at a global MAF of 1,48% at the Genome Aggregation Database (gnomAD-v4.1.0), 226 homozygotes reported, and it has previously been described as a LOF variant associated with lower LDL-C levels (17).

The **APOB c.2630C>T** variant consists of a citidine to timine substitucion that predicts the replacement of an amino acid of proline to leucine at position 877 of the ApoB-100 protein. It is present at a global MAF of 0,04789% at the Genome Aggregation Database (gnomAD-v4.1.0), three homozygotes reported, and it is classified as a VUS in ClinVar. In silico predictors are not conclusive towards the pathogenicity of the variant, that is is located within the N-terminal βα1 domain of ApoB-100, a region not directly involved in the canonical LDL receptor (LDLR)–binding domain

Following the initial findings in the index case, genetic analysis was extended to available family members to better characterize the identified variants and to carry out cascade screening.

### 3.3 Segregation analysis

The index case (III.9) had three offspring (IV.3, IV.4 and IV.5). The eldest one, IV.3 carried the same genotype as her father and was clinically diagnosed of FH. The youngest one, IV.5, did not carried the *LDLR* variant but carried the *APOB* c.2630C>T p.(Pro877Leu) and the *PCSK9* c.137G>T p.(Arg46Leu) variants in heterozygosity, she showed normal LDLc. Interestingly, the individual IV.4 inhered *LDLR* c.2479G>A p.Val827Ile, his lipid levels were within the normal range. Additionally, he showed two more variants, both in *PCSK9* c.137G>T p.(Arg46Leu) in homozygous and the very rare variant, c.2023delG p.(Val675Leufs*130) in heterozygous.

Three relatives of the index case (III.10-12) did not carried the *LDLR* variant and showed normal lipid profile. Two of them carried the *APOB* c.2630C>T p.(Pro877Leu), suggesting that this variant may be not related to FH and it is probably benign. The individual II.1 had hypercholesterolemia and died prematurely due to IAM in his sixties. Molecular analysis of his offspring revealed that III.1 and III.3, both diagnosed with hypercholesterolemia, carried the *LDLR* c.2479G>A p.(Val827Ile), *PCSK9* c.137G>T p.(Arg46Leu) and *APOB* c.2630C>T p.(Pro877Leu) variants in heterozygosity. III.2 had normal lipid profile and only carried the *PCSK9 c.137G>T* p.(Arg46Leu) in heterozygosity. The individual II.2 carried *LDLR* c.2479G>A p.(Val827Ile), *PCSK9* c.137G>T p.(Arg46Leu) and *APOB* c.2630C>T p.(Pro877Leu) in heterozygous and had hypercholesterolemia. Her descendant, III.4 inherited the LDLR variant and showed hypercholesterolemia. The complete familial pedigree is shown in **Figure 1**.

**Figure 1.**
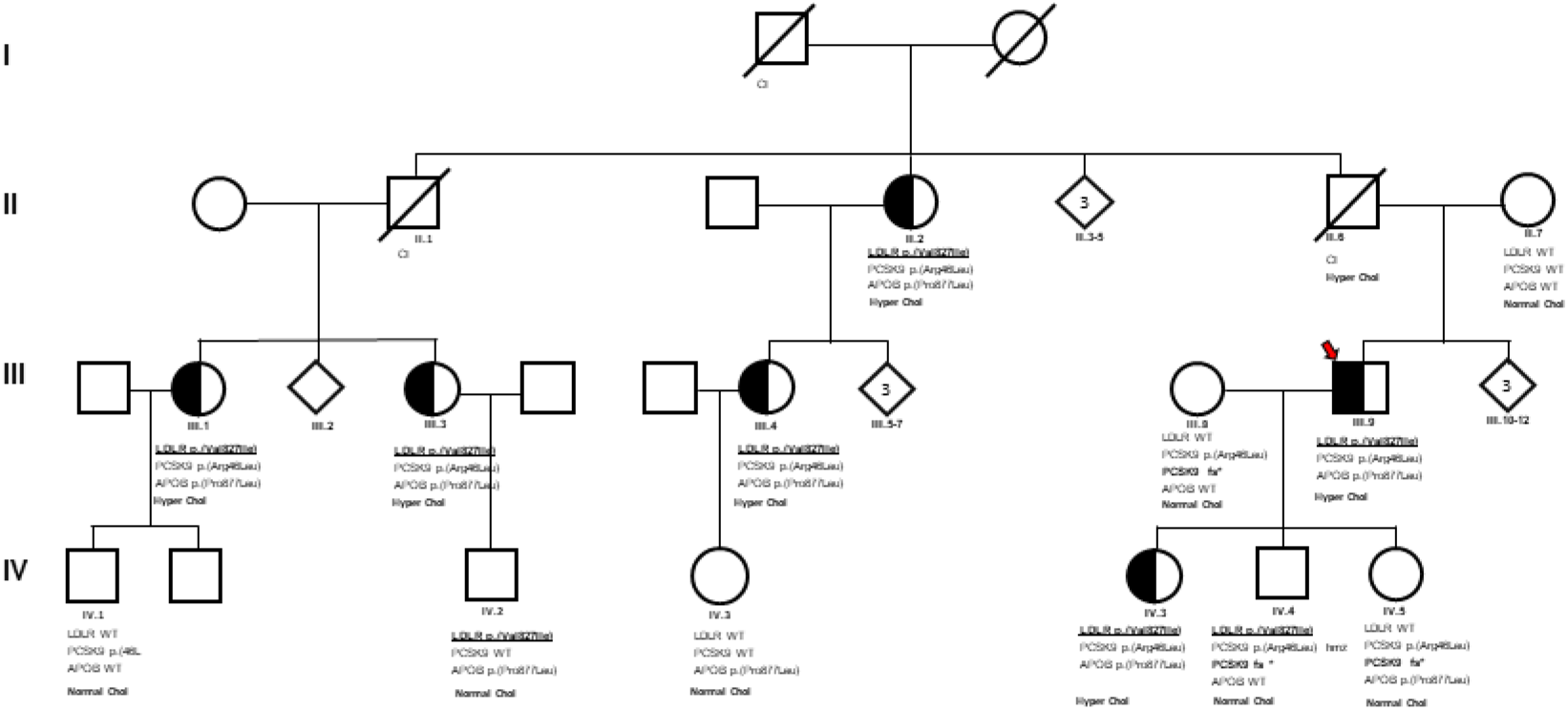
Pedigree of the family showing segregation of *LDLR* and *PCSK9* variants. Squares represent males, circles are females and diamond symbols are used to represent family members without specifying their sex. Individuals fulfilling the clinical diagnosis criteria for familial hypercholesterolemia (FH) are indicated by half-filled symbols. The first row below each symbol correspond to the individual ID following by the genotype at *LDLR, PCSK9* and *APOB*..

Overall, pedigree analysis supports cosegregation of the *LDLR* c.2479G>A variant with the disease, except in two individuals, IV.4 and IV.2, who exhibited normal cholesterol levels despite carrying *LDLR* variant. As we mention above the individual IV.4 carried two additional variants in *PCSK9*, one of which was extremely rare. The in silico characterization of this variant is described below.

### 3.4 Functional characterization of the *LDLR* c.2479G>A p.(Val827Ile)

#### Functional activity

A significant reduction in LDL uptake was observed by flow cytometry in cells expressing the mutant LDLR compared with the wild-type (74% of wild-type), whereas no differences were detected in LDL binding or LDLR expression (**Figure 2**). The results showed that LDLR variant exhibits a moderate functional defect.

**Figure 2:**
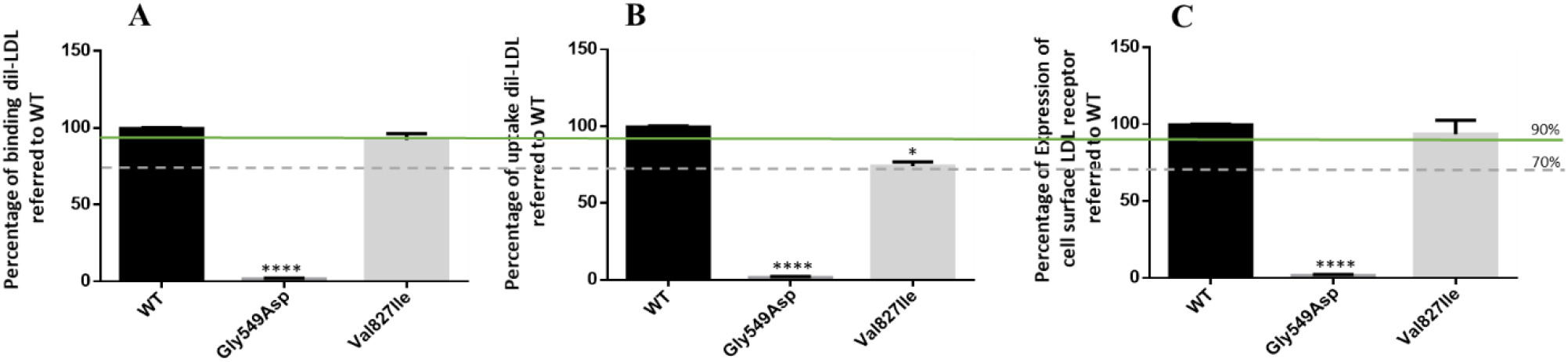
Functional characterization of *LDLR c.2479G>A*; (A) Flow cytometry histogram overlay showing DiI-LDL binding; (B) Quantification of LDL uptake relative to WT; (C) LDLR expression. The upper line represents the benignity cutoff (90%) established by the ACMG/ClinGen guidelines for functional studies. * <0.05; **** <0.0001.

### 3.5 In silico analysis of *PCSK9 c.2023del* p.(Val675Leufs*130)

The variant *PCSK9* c.2023del p.(Val675Leufs*130) consists of a single-nucleotide deletion of a guanine that change the reading frame of the gene extending the translation of PCSK9 by 130 aminoacids beyond the reference stop codon (130/692 ~18%). It is present at a global MAF of 0,0003099% at the Genome Aggregation Database (gnomAD-v4.1.0), no homozygotes reported. It is classified in ClinVar Database as VUS by three submitters (February 2026) and to our knowledge, it has not previously been reported in individuals affected with PCSK9-related disorders in the literature.

This variant affects the C-terminal region of PCSK9, also known as V domain or C-terminal Cys/His-rich domain (CHRD), which is required for PCSK9-mediated degradation of LDLR(18). This region has a cylindrical shape and consists of three subunits built from six antiparallel β strands connected by internal disfulfide bonds that ensure the proper folding of the protein (Figure 3). It also contains a large number of histidines, which may help to increase PCSK9’s affinity for LDLR at endosomes’ low pH(19).

**Figure 3.**
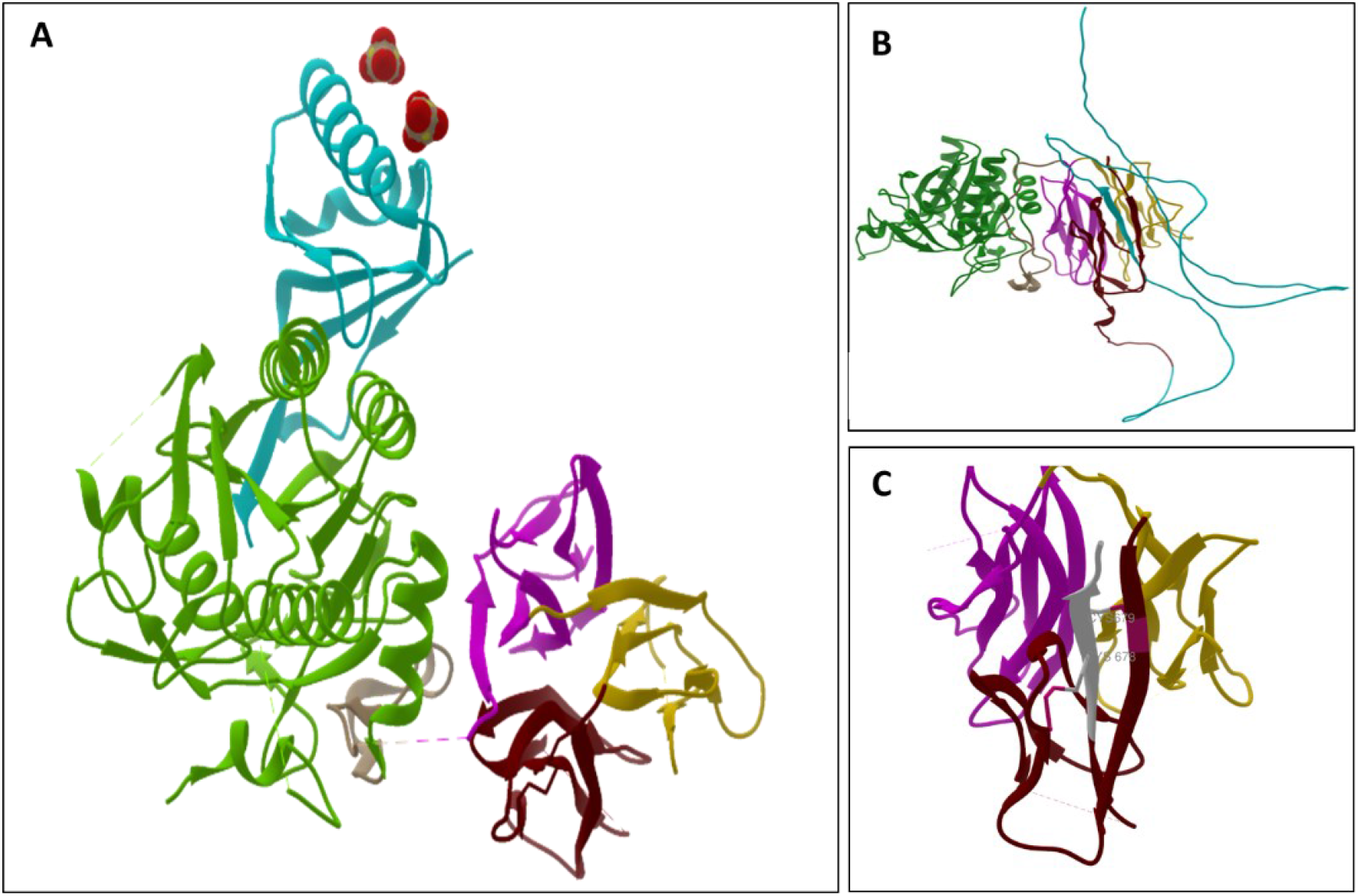
PCSK9 model. A. Structure of native PCSK9 protein (PBD:2PMVW). PCSK9 comprises a prodomain (light blue, aa 31-152), a catalytic domain (green, aa 153-421) and a CHRD domain (aa 453-692) that is composed of three modules. M1: pink, aa 453-529; M2: yellow, aa530-603; M3: red, aa 604-692). B. Structure of mutant PCSK9 protein (Alphafold). The mutated model predicts an extended protein that fails to form the terminal β-sheet present in the wild-type structure, resulting in loss of the native C-terminal folding. C. Loss of Cys678 and Cys679. The mutant sequence is showed in white. Lost residues Cys678 and Cys679 are labeled and the loss of both disulfind bonds is showed in dark pink

In silico structural analysis of p.(Val675Leufs*130) predicts an altered protein that exhibits a marked increase in structual disorder at the C-terminal domain (**Figure 3**) and the loss of Cys678 and Cys679, two highly conserved residues of the subdomain 3. Both cysteines are critical in the beta-sheet stabilisation: Cys678 from strand strand 6 forms a disulfide bond with Cys626 from β strand 2 whereas Cys679 from β strand 6 forms a disfulfide bond with Cys608 from β strand 1

Loss of terminal cysteines of the C-terminal domain has previously been reported as a cause of aberrant folding of PCSK9. Zhao et al (2006) proved that variant p.(Cys679X) caused aberrant folding of the protein, as evidenced by abnormal migration patterns on SDS-PAGE gel after treatment with dithiothreitol (DTT). Furthermore, expression studies in HEK-293 cells revealed that the Cys679X variant impairs PCSK9 secretion by retaining the abnormal protein in the endoplasmic reticulum (ER)(20). Taking this information into consideration, together with the predicted destabilization caused by loss of the sheet-like native conformation of subdomain 3 and steric hindrance from protein elongation, as supported by the structural model, we suggest the p.(Val675Leufs*130) variant reported in this study is likely to have the same effect.

### 3.6 Serum PCSK9 levels

Quantitative analysis of serum PCSK9 was performed using an ELISA assay. The assay included serum samples from the son (IV.4), additional family members and a subset of normolipidaemic controls. We found that individual IV.4 carrying the *PCSK9*:[c.137G>T;c.2023delG]+[c.137G>T] had markedly lower circulating PCSK9 protein levels compared with other family members and the control group (**Figure 4**). This reduction is consistent with a loss-of-function or hypomorphic effect of the *PCSK9* variant.

**Figure 4.**
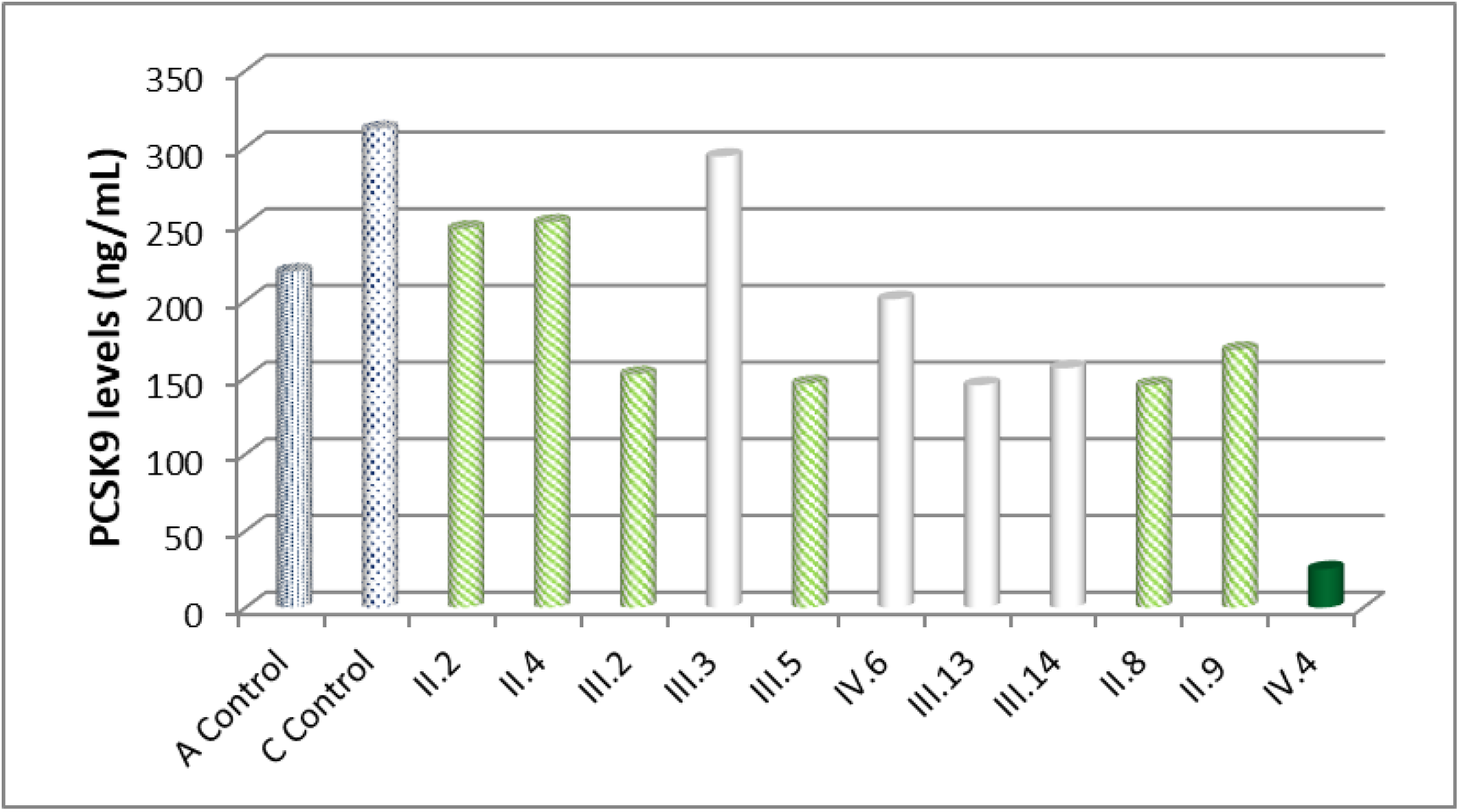
Quantitative analysis of serum PCSK9 levels (ng/mL). Numbers correspond to family members according with figure 1. Dashed bars indicate adult controls (A Control) and child controls (C Control). Hatched bars indicate individuals carrying the *PCSK9* c.137G>T, whereas empty bars indicate individuals without *PCSK9* variants. The solid bar corrresponds to individual IV.4, who carried *PCSK9* [c.137G>T;c.2023delG]+[c.137G>T].

### 3.6 Classification of *LDLR* c.2479G>A variant integrating available data

The combined segregation data and functional analyses, which demonstrated a moderate reduction in LDLR activity in our in vitro model, support an association between the LDLR variant and FH phenotype. Although the variant does not fully satisfy all criteria of the ACMG/ClinGen FH-specific LDLR curation guidelines, the overall body of evidence is consistent with its involvement in the FH phenotype observed in this family. The *LDLR* c.2479G>A variant co-segregates with hypercholesterolemia across multiple affected branches of the pedigree. The absence of an FH phenotype in the proband’s son may be explained by a protective effect conferred by the *PCSK9* genotype. Collectively, the extended segregation data, supportive in silico pathogenicity predictions (REVEL score = 0.736), and the observed moderate functional impairment support an association between this variant the FH phenotype.

## Discussion

Herein, we describe a family carrying the longstanding controversial *LDLR* variant, c.2479G>A p.(Val827Ile) which has undergone multiple reclassifications over time. This variant had been previously reported associated with FH and was historically referred to as “FH New York-5”(21). Current public database evidence remains heterogeneous: ClinVar aggregates conflicting interpretations across submitters, and a substantial fraction of laboratories still classify the variant as a VUS (22). According to ClinGen FH Variant Curation Expert Panel LDLR-specific ACMG/AMP specifications, this variant has also been evaluated as VUS, typically supported by limited segregation, computational evidence and high population frequency (e.g., PP1/PP3 depending on dataset strength) (23).

In large reference datasets, LDLR c.2479G>A p.(Val827Ile) variant is rare, with a global MAF of 0,046%, but it is markedly enriched in specific genetic ancestry groups such as Ashkenazi Jews (gnomAD subpopulation frequency reported at ~0.018, with homozygotes)(24). The high prevalence of this variant may reflect a founder effect, consistent with studies highlighting that some Jewish subpopulations exhibit an FH prevalence 7 times higher than that of general Caucasian populations. This fact illustrate why allele-frequency evidence for p.(Val827Ile) should be interpreted in the context of ancestry and local FH epidemiology rather than using global prevalence assumptions alone(25).

Moreover, the reported functional consequences of this variant vary across studies, further contributing to uncertainty regarding its pathogenicity. In our CHO-ldlA7 model, LDLR p.(Val827Ile) retained ~74% of wild-type LDL uptake consistent with a borderline internalization defect rather than a severe loss of function. This data is consistent with a borderline internalization defect and align with previous studies reporting either mild impairment to near-normal behaviour depending on assay design and cellular context. For example, a systematic cell-based pheotyping study evaluating rare LDLR missense alleles reported that many variants exhibit context-dependent functional penetrance, and functional readouts can refine cardiovascular risk inference in cohorts(26). More recently, massively parallel functional mapping of nearly all LDLR missense substitutions generated quantitative sequence–function maps for both LDLR abundance and LDL uptake, providing an additional framework in which some conservative substitutions may score near-normal in specific heterologous assay settings(27).

The c.2479G>A p.(Val827Ile) substitution lies within the FxNPxY internalization motif in the LDLR cytosolic tail, a signal required for efficient packaging of LDLR into clathrin-coated pits through binding to PTB-domain endocytic adaptors (5). Structural studies have shown that the ARH/LDLRAP1 PTB domain recognizes not only the core FxNPxY residues but also an extended LDLR tail segment (I(−7)xF(−5)xNPxY(0)QK(+2)) that adopts a distinctive “hook-like” conformation, providing a plausible mechanistic basis by which even conservative substitutions at non-core positions could modulate adaptor engagement and endocytic efficiency(28). Conceptually, variants in the LDLR cytosolic domain can produce class 4 LDLR defects, characterized by preserved receptor surface expression but impairment internalization(6).

An explantion for these discrepancies is the tissue-specific hierarchy and redundancy of LDLR endocytic adaptors. Genetic and cellular studies established that ARH/LDLRAP1 has a tissue-restricted, physiologically critical role in hepatic LDL clearance, whereas LDLR internalization can be maintained in other cell types through alternative adaptor proteins(9). In non-hepatic contexts, Dab2 can act as a major LDLR sorting adaptor (including HeLa cells and fibroblasts), and in certain settings can function independently of AP-2 and ARH(29). Moreover, mechanistic dissection of clathrin-mediated uptake demonstrated that LDLR’s FxNPxY-type signal is recognized redundantly by Dab2 and ARH, and that simultaneous depletion of both adaptors leads to marked LDLR surface accumulation. Importantly, Dab2 expression is exceptionally low in hepatocytes, which explains why ARH loss produces severe hypercholesterolemia in vivo(30). CHO-ldlA7 cells (and other non-hepatic models) may systematically underestimate the functional impact of subtle NPxY-motif variants that primary disrupt ARH-dependent endocytic sorting in hepatocytes, leading to false negative or borderline in vitro readouts despite clinically relevant impairment of hepatic LDL clearance. Therefore, given the adaptor redundancy outside the liver and the dominant role of ARH in hepatic LDL clearance, functional interpretation of NPxY-motif variants should prioritize context-aware models rather than relying on a single heterologous system as definitive evidence.

In summary, our data support a modest but consistent functional impact of the *LDLR* c.2479G>A variant in CHO-ldlA7 cells, and provide an explanation for the differences in retained LDLR activity reported across studies. Functional studies along with the results of the segregation analysis in family members demonstrate that the LDLR c.2479G>A variant cosegregates with the phenotype and supports the involvement of the in the FH phenotype observed in this family.

Case IV.4 represent the only exception in the segregation analysis, given that he carried the c.2479G>A variant but was normolipidaemic. The prevalence of normolipidaemic individuals who have molecular diagnostic of hypercholesterolemia has been reported to be low (1,6%)(31). This low prevalence is biologically plausible because penetrance of *LDLR, APOB or PCSK9* variants is generally high but incomplete, and LDL-C shows wide inter-individual variability even among carriers(32) (22). Variant type also influences phenotypic severity: LDLR null/receptor-negative variants are typically associate with higher LDL-C and more severe clinical manifestations than receptor-defective variants, which generally present with milder phenotypes(33). In addition, protective modifiers can blunt the biochemical phenotype, including co-inheritance of LDL-lowering variants (e.g., loss-of-function in *APOB* or *PCSK9* (17), a favourable polygenic background (low burden of LDL-C-raising alleles)(34), and lifestyle factors that can modulate risk and, to some extent, lipid levels(35).

In this case, the absence of hypercholesterolemia, despite carrying the family variant, can be plausibly explained by the concomitant presence of LOF variants in *PCSK9* c.137G>T p.(Arg46Leu) *in homozygous and* c.2023delG p.(Val675Leufs*130) in heterozygous. The PCSK9 p.(Arg46Leu) variant is a well-established LOF variant that has been consistently associated with lower LDL-cholesterol concentrations, cardiovascular disease and reduced circulating PCSK9 levels (17) (36) (37). This variant has been associated with an allele-dose effect, and immunoassay-based studies have shown that (heterozygous vs homozygous), studies measuring circulating protein by immunoassay show that p.(Arg46Leu) carriers display significantly lower circulating PCSK9 concentrations, supporting a stronge biological impact as the number of p.(Arg46Leu) alleles increases(38). In our patient, the co-ocurrence of an adictional *PCSK9* variant, likely further reduces functional PCSK9 availability, providing a plausible explanation for the markedly low serum PCSK9 levels observed. Mechanistically, this combined genetic background would be expected to attenuate LDLR degradation and promote LDLR recycling, thereby contributing to normal LDL-cholesterol concentrations despite the presence of an FH-causing variant. Overall, the coesxitence of p.(Arg46Leu) and p.(Val675Leufs*130) is consistent with an additive protective modifier effect that may counterbalance the anticipated LDL-c elevation.

## Conclusion

In conclusion, while cascade testing of the familial pathogenic variant in first-degree relatives remains the recommended and cost-effective strategy for cardiovascular prevention, our findings support extending genetic analysis—particularly when the lipid phenotype is discordant with the expected genotype. Broader NGS-based approaches can uncover modifier variants and gene–gene interactions that help explain the clinical heterogeneity of familial hypercholesterolemia and refine risk estimation, since protective alleles present in an apparently unaffected carrier may not segregate to descendants, potentially unmasking hypercholesterolemia. This case exemplifies the interpretative challenges introduced by extensive sequencing and underscores the need to integrate clinical, biochemical, genomic, and functional evidence to reach robust conclusions. Collectively, the data indicate that PCSK9 can act as a disease modifier by attenuating the phenotypic impact of an LDLR pathogenic variant, reinforcing the relevance of considering genetic interactions to improve counselling and advance precision medicine in dyslipidaemia.

## Data Availability

All data produced in the present study are available upon reasonable request to the authors

## Acknowledgements

We sincerely thank all members of the family for their generous participation in this study.

